# Can You Hear What I Hear: Exploring ability and perspective when matching loudness of auditory verbal hallucinations to audio volume

**DOI:** 10.64898/2026.07.27.26359058

**Authors:** James Heap, R Brooke Stephenson, Clare L Beasley

## Abstract

**Introduction:** Auditory verbal hallucinations (AVH) affect 60-80% of people with schizophrenia, yet existing assessment tools inadequately capture their phenomenological complexity.

**Aim:** To determine whether individuals with schizophrenia spectrum disorders could accurately match auditory verbal hallucination loudness to an external audio track, and to explore participant perspectives on this approach.

**Methods:** Eight participants with schizophrenia spectrum disorders and active AVHs rated loudness via a Likert scale and by adjusting a headphone audio track to match their experience. Structured interviews and thematic analysis captured participant viewpoints.

**Results:** No significant correlation was found between audio tool and Likert scale scores. Seven of eight participants reported the audio tool provided greater precision in quantifying AVH loudness and better enabled them to convey their internal experience to others.

**Discussion:** Audio-matching tools may offer meaningful advantages over traditional scales for quantifying AVH loudness, even where statistical convergence with existing measures is absent.

**Limitations:** Small sample size; loudness alone cannot fully capture the qualitative experience of hearing voices.

**Implications:** This tool shows promise for longitudinal tracking of AVH loudness.

**Recommendations:** Further investigation of digital approaches to AVH assessment is warranted.

**Relevance Statement:** Mental health nurses are often the clinicians who assess and monitor auditory verbal hallucinations, yet they rely on rating scales that ask patients to score or describe their voices. This study explores an approach that enables patients to demonstrate the loudness of their voices rather than describe it. For nursing practice, it may offer a concrete, patient-led way to quantify symptom severity, track change over time, and communicate that experience with others. With development and validation, the approach may support improved clinical assessment, strengthen rapport and inform how mental health nurses evaluate and plan care for people with psychosis.

**Accessible Summary:** *What is known on the subject:*

- Many people living with psychotic conditions such as schizophrenia hear voices that others cannot hear, known as auditory verbal hallucinations (AVH).
- Voices vary in how loud they seem, and louder voices can be more distressing.
- Clinicians usually measure loudness using rating scales that ask people to score their voice hearing experiences.
- Prior research has called for the development of new approaches in the area of AVH assessment.

*Originality:*

- This study tested a new approach: participants adjusted the volume of a sound played through headphones until it matched how loud their voices were.

*Significance:*

- Most participants (seven of eight) felt the headphone method captured their experience better than a rating scale and helped them explain it to others.
- The tool could help mental health nurses and other clinicians track whether voices are getting louder or quieter over time, including in response to treatment.
- Larger studies are needed, but this study suggests that simple technology could improve how voice-hearing is understood and discussed.

## Introduction

Auditory hallucinations that are experienced as voices are referred to clinically as auditory verbal hallucinations (AVHs) (de Leede-Smith & Barkus, 2013). AVHs are typically considered to represent internally generated events that are not recognised as self-generated and are misattributed to an external source. Considered one of the key symptoms of psychotic disorders, AVHs are present in around 60-80% of people with schizophrenia, but can also occur in other psychiatric disorders such as bipolar disorder, substance use disorder, dissociative disorder and major depressive disorder (Steenhuis et al., 2019; Waters, 2010; Lim et al., 2016).

AVHs possess complex phenomenological characteristics which include loudness; localization; explanation of origin; voices speaking in third person; controllability; number of different voices; frequency; duration; types of voices experienced; disturbances to daily functioning and emotional valence of the voices (de Leede-Smith & Barkus, 2013). Patient experiences comprise varied combinations of these characteristics, demonstrating significant interindividual variability in form and content. Given this heterogeneity, understanding and quantifying an individual patients AVHs presents a challenge that calls for a variety of approaches and perspectives (Jardri et al., 2019). AVH assessment largely relies on self-report, and features of the hallucinatory experience may be difficult for a patient to articulate to clinicians. While validated scales, such as the Psychotic Symptom Rating Scales (PSYRATS; Haddock et al., 1999) can be used to assess AVHs, these scales are often interviewer administered, lengthy and subjective. As such, the development of new scales, and the refinement of those already in existence, is warranted (Ratcliff et al., 2010). Moreover, increased involvement of voice hearing consumers in the development and testing of existing and novel assessment tools is imperative (Ratcliff et al., 2010).

One important aspect of AVH is loudness. While AVHs are often perceived at a normal conversational volume, voices that are soft/whispering, loud, yelling or screaming may also be experienced (McCarthy-Jones et al., 2012). Greater loudness of AVH among patients with schizophrenia has been associated with distress and disruption to life (Kim et al., 2019). While currently typically measured using subjective rating scales, AVH loudness may be amenable to a finer subcategorization, which may impart important clinical information. As such, the aim of this study was to develop a novel audio tool to quantify loudness of AVHs in subjects with schizophrenia spectrum disorder (SSD) and to explore patient perspectives on the use of this approach to better understand their personal experience of AVHs.

## METHODS

### Ethics

The study was approved by UBC Children’s and Women’s Research Ethics Board (certificate H21-03500) and the BC Mental Health and Substance Use Services Research Committee. Written informed consent was obtained from each patient prior to enrollment in the study. The study was conducted in accordance with the Tri-Council Policy Statement: Ethical Conduct for Research Involving Humans (TCPS 2), the Canadian national standard, which incorporates the principles of the Declaration of Helsinki.

### Participants

Participants aged 19 and older with a self-reported diagnosis of schizophrenia or other psychotic disorder, who were actively experiencing AVHs and who were willing and able to provide written informed consent were recruited through community mental health resources in British Columbia, Forensic Psychiatric Services regional clinics and the Forensic Psychiatric Hospital. Exclusion criteria included self-report of methamphetamine use within the last week and use of hearing aids or hearing impairment.

### Quantitative Assessment of AVH Loudness

Participants were first asked to rate the loudness of their current AVHs against a 7-point author-created paper-based Likert scale (0 = no speaking at all; 1= a whisper; 2 = quiet talking; 3 = normal talking; 4 = loud talking; 5 = shouting/yelling; 6 = unbearably loud screaming).

Participants then donned a pair of over-the-ear noise cancelling headphones (HD-450BT, Sennheiser) connected to a smartphone (iPhone SE, Apple) via Bluetooth. An audio track (a recording of the lead researcher reading a bread recipe in the French language, recorded using the Voice Memos app, and played in reverse) was then played through the headphones. Participants were asked to adjust the volume of the smartphone upwards to a loudness that matched their AVHs. The final volume setting of the smartphone was recorded. The sound pressure level associated with each discrete volume step of the smartphone-headphone configuration was quantified and converted to decibels (Supplementary figure 1).

### Statistical analysis

The relationship between AVH loudness described using the Likert scale and matched to the audio track was determined using Spearman’s Rank correlation (Graphpad Prism 10 for MacOS).

### Qualitative Interviews

Interviews were conducted by trained researchers who had experience working with individuals with mental illness. Each participant was asked five questions (Table 1). All interviews were transcribed verbatim and examined using thematic analysis.

**Table 1.**
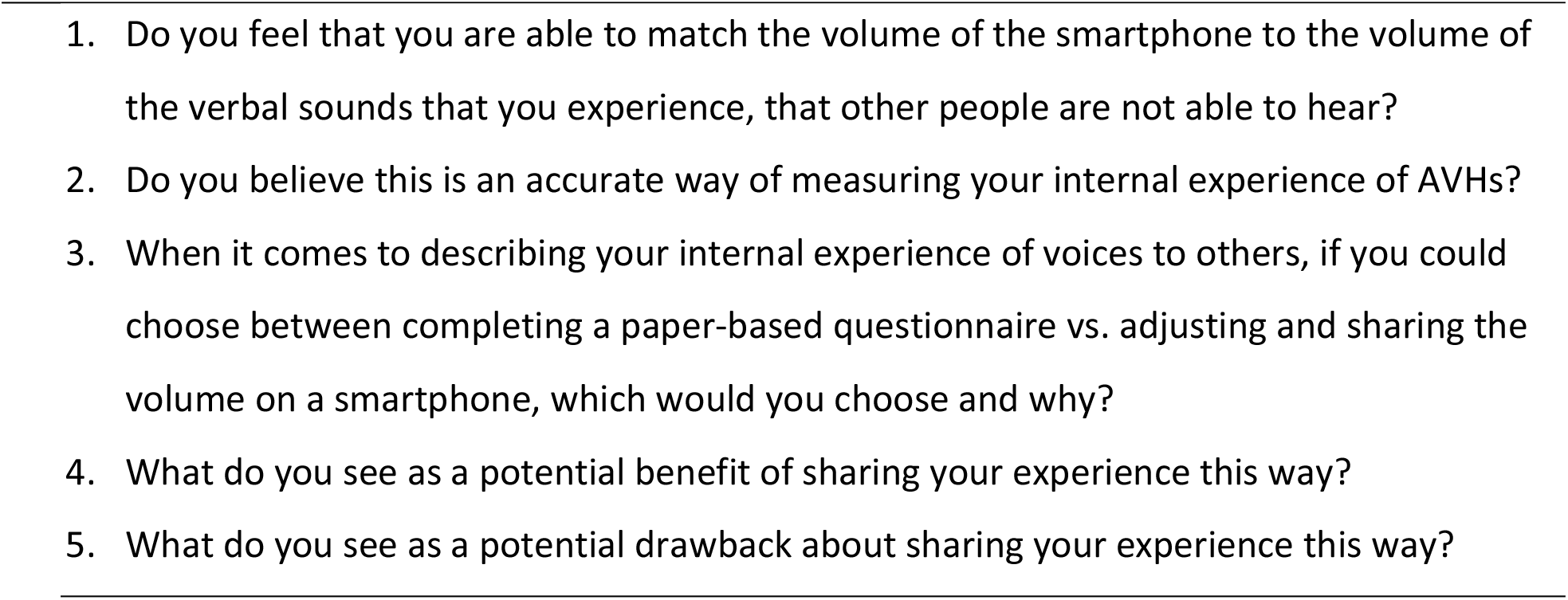

## RESULTS

### Participants

A total of eight individuals participated in the study. Three participants reported a diagnosis of schizoaffective disorder and five reported a diagnosis of schizophrenia. Three participants identified as female and five as male, with all identifying as cisgender. Participants were aged between 38 and 72 years, with a median age of 50.8 years. Five participants identified as White, two as Chinese and one as Southeast Asian.

### Quantitative Assessment of AVH Loudness

Ratings of AVH loudness on the paper based 7-point Likert scale ranged from 1 (whisper) to 3.5 (normal to loud talking), with median value of 2.5 (quiet to normal talking). Ratings of AVH loudness using the audio tool ranged from 3/16 (approx. 39dB) to 11/16 (approx. 72dB), with a median of 4.75 (approx. 48dB) (Figure 1). Although generally in agreement, two participants provided values that differed substantially between measures; one reported experiencing their voices as ‘quiet talking’ according to the Likert scale, but exceeded 70dB using the audio tool, equivalent to a group conversation, a vacuum cleaner or an alarm clock (American Speech-Language-Hearing Association, 2025). Conversely, one participant reported experiencing their voices as ‘normal talking’ according to the Likert scale, but recorded 39dB using the audio tool, equivalent to a quiet room. Ratings obtained using the two methods were not significantly correlated (r=0.279, p=0.491).

**Figure 1.**
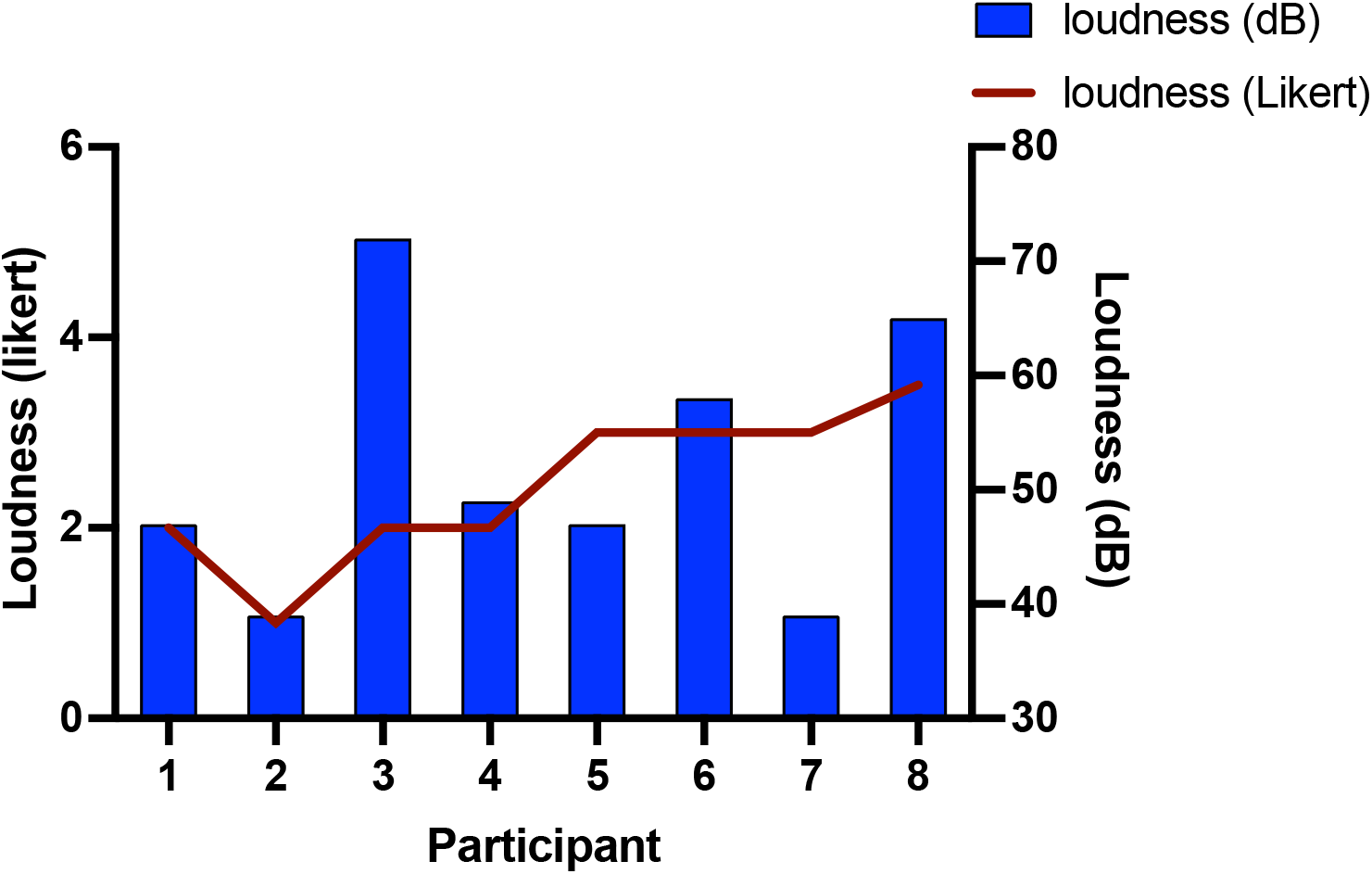
Comparison of Likert scale ratings and smartphone volume matching (dB)

## Themes from Qualitative Responses

Analysis of the qualitative responses highlighted several key themes:

### Theme 1: Participants felt that they were able to accurately match their AVHs with the audio volume

All participants were able to match the loudness of their AVHs to the loudness of the audio tool volume. Seven out of eight study participants reported the audio tool provided an accurate method for describing the loudness of their AVHs, noting *“…the volume of the voices can change but that is the level of the voices today”* - (participant 6503). Two participants reported that the loudness of their AVH fell between two volume steps. One participant was unsure whether the audio tool accurately represented the volume of their AVHs but responded “*I think so*”.

### Theme 2: Preference for smartphone method over a paper-based method

Seven out of eight participants voiced a preference for the smartphone audio tool method, describing it as ‘*more live…more relevant…more real’* – (participant 6501) and *‘more effective’* – (participant 6502). One participant noted that “…*it’s difficult to put it on paper*…*it’s not always true*…*but if you hear the voice and can adjust the volume then you know you’re doing the right thing*” - (participant 6505). Participants felt that the smartphone audio tool was more accurate than the Likert scale, commenting that “*It’s participant confirmed, allowing for more accuracy*” - (participant 6500), “*I think it’s more like an accurate way of describing of how loud my voices are*” - (participant 6502), and “…*This is an actual technical rating whereas a paper-based rating is just guess work*” – (participant 6500). One participant was not in favour of either method, stating *“I don’t like either one…” – (participant 6506)*.

### Theme 3: Potential benefits of a smartphone audio track volume matching method

Several participants responded that the smartphone method offered a mechanism for conveying their internal AVH experience to others and thus had potential to promote empathy as well as influence clinical care: “*It would help family members understand*…*help others understand what’s going on for me*” - (participant 6504), “*Empathy*…*understanding*…*I’m not saying you can completely ‘get’ schizophrenia by sharing headphones with someone, but it gives you a quick idea of what it’s like to have schizophrenia*” - (participant 6507) “*…I can make it real for the other person in terms of what I am hearing*…*because you can take the sounds and you can show somebody the level of sounds of the voices*…*it’s a real way to show somebody what it’s like to have schizophrenia*” – (participant 6503); “*It could give psychiatrists some knowledge about auditory hallucinations. They could treat me better with medicine and build empathy*” - (participant 6501).

### Theme 4: Potential drawbacks to the smartphone volume matching method

Some participants noted potential drawbacks and concerns regarding the smartphone audio volume matching method. These included:

#### 4.1 Concerns about the inability of the smartphone method to fully capture the complexity of the AVH experience

Several participants highlighted the importance of considering additional phenomenological characteristics of AVHs, such as the quality and diversity of the voices and the variability over time. *“What they’re saying*…*the intensity, and how to react to the voices*… *but they used to be really dark*…*the quality of the voices*…*but this is one way to approach it that helps*” - (participant 6504); “*I have ambient sound hallucinations, disembodied hallucinations and TV hallucinations*…*the TV hallucinations can be just as loud as the TV*…*the loudest are when people are talking to me and are just as loud as human voices*…*these ones are super real*.” – (participant 6505); “*Because the loudness varies*…*depending on when the encounter happens it may influence the rating scale*” - (participant 6501). In addition, dissimilarity between listening to the audio track and internally perceived voices was noted. “*The loudness resembles it*…*but it’s different*…*It’s not the same thing, it’s analogous but it’s not the same thing*…*One reason is because it comes from outside my head, but my hallucinations originate from inside my head*…*yes I can match the loudness, but I can tell the difference between the voices coming from outside*.” – (participant 6507).

#### 4.2 Concerns about how the information might be used

One participant voiced concerns over sharing the loudness of their AVHs with clinicians, noting “*Maybe the doctor would see it and overmedicate me*” - (participant 6501). Participants also expressed apprehension that the audio method might increase stigma, and could potentially cause fear and discomfort in others. “*Maybe schizophrenia would be stigmatized more*…*”* - (participant 6501); *“I don’t know*…*it might inflict fear or curiosity in them*…*sometimes the voices are very unfriendly or vehemently influencing your piece of mind*…*if you were listening to angry or scary voices … that might be scary to people listening”* (Participant 6507).

## Discussion

The results from this study suggest that seven out of eight participants were able to accurately match the loudness of their AVHs to an audio track played on a smartphone. Some participants chose a number between two available volume steps, suggesting that the tool offered the opportunity to describe the loudness of their AVHs with much greater precision than offered by the Likert scale. The median values provided by participants using the Likert scale (quiet to normal talking) and smartphone audio tracks (48dB, equivalent to moderate rainfall) were comparable, suggesting that most participants experienced AVHs at a volume similar to a quiet to typical conversation. The values recorded using the two methods were not correlated, which may be due to two participants reporting divergent scores for the Likert and audio scales. This raises the question whether a smartphone audio volume matching method could shed light on an internal experience that is different than what is being described verbally.

In general, participants reported the smartphone audio tool was more accurate than the Likert scale, with participants describing the audio tool as “*more real*,” “*more accurate*,” and “*more relevant*”. Furthermore, the smartphone audio tool may offer a novel approach to helping people describe their internal experience of AVHs to others, including clinicians and family members. Participant responses suggest a smartphone audio tool might have value in promoting understanding and empathy in others, which in turn may have potential to reduce stigma.

We recognise that this study has limitations. First, the sample size of eight participants limits how broadly these findings can be applied. While this size is consistent with recommendations for homogeneous samples with narrowly defined objectives (Kuzel, 1992; Hennink & Kaiser., 2022), larger studies are needed to confirm the results. Further, participant diagnoses were based on self-report rather than structured clinical interviews. Although self-reported diagnoses have shown acceptable validity in voice-hearing research (Woods et al., 2015), future studies may benefit from diagnostic confirmation. We did not control for potential medication effects or assess current symptom severity, nor did we test hearing proficiency. Moreover, we assessed AVHs loudness at a single point in time. Some participants noted that their AVHs loudness varies, and we did not examine responses throughout the same day or on different days.

Second, only one aspect of AVHs was quantified in this study. Several participants highlighted the complexity of their AVH experience, shedding light on the added impact of emotional valence of AVHs, and the variability of their hallucinations over time. The distinction between externally generated sounds and internally perceived voices also raises questions about whether external sound can truly represent an internally perceived experience. Participant experiences of distinct types of hallucinations (e.g., “*ambient*,” “*disembodied*,” “*TV hallucinations*”) and the impact of localization on their experiences highlights variability in the way individuals experience AVHs and the complexity of the AVH experience. However, we suggest that, although an audio volume matching tool focussed only on loudness is unable to capture the complexity of AVHs, it may be useful as part of a suite of tools addressing a wider range of phenomenological characteristics.

Third, AVHs were described using an author-created Likert scale rather than established instruments. Future research should examine how audio volume matching compares to existing measures, such as the PSYRATS, which has a validated loudness item (Haddock et al., 1999). Our study also assumes that participants had insight into the hallucinatory nature of their AVHs, which we described as “voices that no one else around you can hear”.

## Conclusion

This study revealed that individuals diagnosed with a schizophrenia spectrum disorder were able to match the loudness of their AVHs to the volume of an audio track playing via a smartphone device. Most participants described the approach as being an accurate means of measuring the loudness of their internal experience of AVHs and highlighted possible benefits of using this approach in helping to convey their internal experience to others, noting a potential for building empathy through improved understanding. The audio tool has the potential to be useful in clinical settings, for example shedding light on treatment efficacy. However, AVH loudness is not constant, and further research is required to examine how volume differs over time and the relationship with symptom severity. Limitations of the tool in failing to capture complex phenomenological characteristics beyond loudness alone were noted. Overall, we suggest that the use of audio matching tools is worthy of further exploration in this context.

## Data Availability

All data produced in the present work are contained in the manuscript

## Author Contributions

JH conceived and designed the study, prepared the study proposal and research ethics application, performed the data collection and analysis, and drafted the manuscript. CLB provided research mentorship and supervision, contributed to the development of the study’s ideas, participated in data analysis, and revised the manuscript. RBS contributed to the development of the study’s ideas, participated in recruitment and data collection and analysis, and critically reviewed and revised the manuscript. All authors gave final approval of the version to be published and agree to be accountable for all aspects of the work.

## Acknowledgements

We thank Will Howie (UBC School of Music) for his expertise in measuring headphone output levels. This study was conducted as patient-oriented research with the involvement of a patient partner who contributed to review of the study plan and to data collection, and who wishes to remain anonymous. We are grateful for their contribution.

