## Supplementary Figure 1 for "Can You Hear What I Hear: Exploring ability and perspective when matching loudness of auditory verbal hallucinations to audio volume"

**Supplementary Figure 1. Smartphone volume step vs sound pressure level (dBA)**

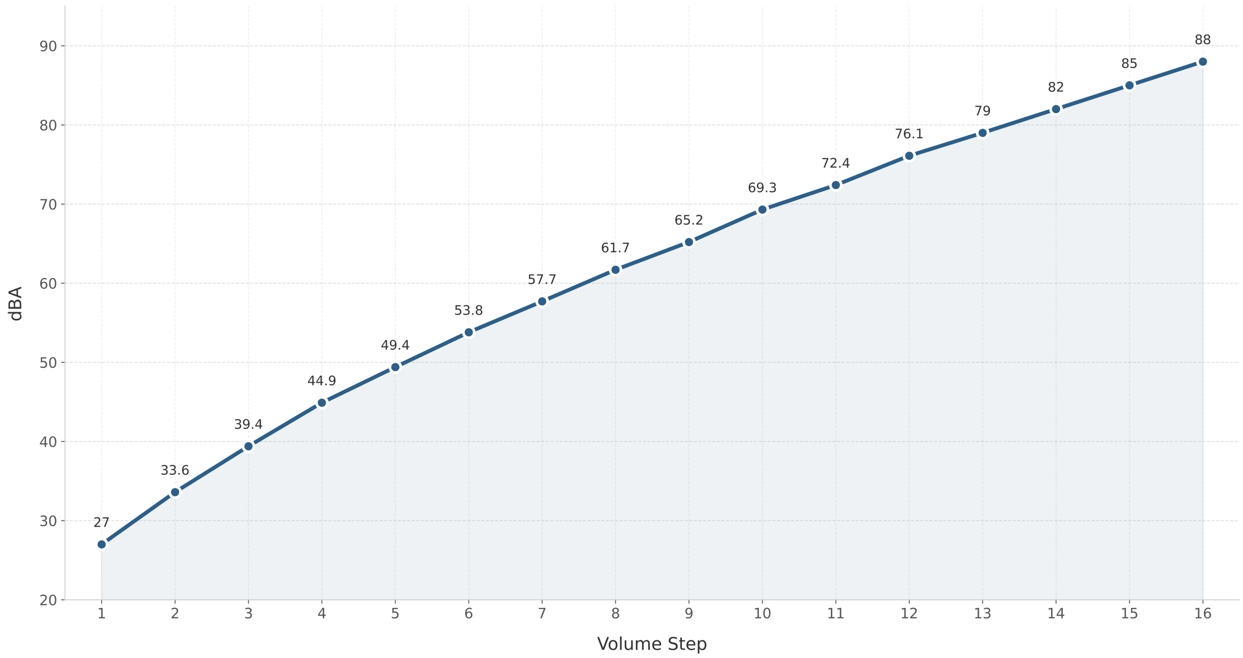


*dBA - Approximate average measured sound pressure level (dBA) of the left headphone, at the ear
